# “Catechol-O-methyltransferase gene Val158Met polymorphism and prostate cancersusceptibility”

**DOI:** 10.1101/2020.04.16.20067736

**Authors:** Pradeep Kumar, Vandana Rai

## Abstract

Prostate cancer is one of the most common and a serious malignancy of males and it is well reported that estrogen plays a pivotal role in prostate carcinogenesis. Catechol-O - methyltransferase (COMT) catalyzes the inactivation of estrogens. Several studies have investigated the association of COMT gene Val158Metpolymorphism with prostate cancer, but results were inconsistent and inconclusive. Hence, to assess this association, we performed a meta-analysis of all published case-control studies. Pubmed, Springer link, Google Scholar, Elsevier and Springer link databases were searched for case-control studies. Odds ratios (ORs) with 95% confidence intervals (CIs) was used as association measure. Statistical analysis was performed with the software program MIX and MetaAnalyst. In the current meta-analysis, 11 case control studies with 3381 prostate cancer cases and 3,276 healthy controls were considered. The results indicated no significant association between COMT Val158Met polymorphism and prostate cancer risk using allele contrast, co-dominant and homozygote models (allele contrast: OR= 0.92; 95% CI 0.85 to 0.98=; p= 0.02; co-dominant: OR=0.81; 95% CI= 0.85 to1.07; p= 0.46; homozygote: OR= 0.81; 95% CI= 0.70 to 0.95, p= 0.008), but showed significant association with dominant and recessive models (dominant: OR 1.18=; 95% CI= 1.03 to1.34; p= 0.01; recessive: OR= 1.54; 95% CI= 1.1 to 2.07; p = 0.003). In subgroup analysis meta-analysis using recessive genetic model showed significant association between COMT Val 158Met polymorphism and prostate cancer risk in both Asian and Caucasian populations. In conclusion, results of present meta-analysis supports that the COMT Val158Met polymorphism is risk factor for prostate cancer.

## Introduction

Prostate cancer is thesecond most common cancer in men worldwide (Center et al.,2012).Intrepithelialneoplasia, adenocarcinoma androgen-dependent and adenocarcinoma androgen-independent or castration-resistant are three developmental stages of prostate cancer. Although the etiology of prostate cancer remains unknown, but age, ethnicity, family history and steroid hormones appear to play a role (Bosland,2000; Bostwick et al., 2004). Its prevalence is disproportionately high in African population, and is less common in Caucasianand Asian populations (Hsing and Chokkalingam, 2006).There is ample evidence supporting the notion that genetics plays a key role (Kolonel, 1997; Parkin et al., 1997; Schaid, 2004). Estrogens, and/or catechol metabolites, have been identified as potential carcinogens for prostate cancer (Yuen et al.,2005).Various studies have investigated the associations between polymorphisms of genes encoding enzymes involved in estrogen metabolism and the risk of prostate cancer(Cancel-Tassin and Cussenot,2005; Cussenot et al.,2007).

Three major estrogens exist in vivo: estrone (E1), estradiol (E2), and estriol (E3) and these estrogens are metabolized by phase I metabolizing enzymes like cytochrome 450. These enzymes metabolically activate procarcinogens to reactive electrophilic forms, reactive oxygen species (ROS), which can damage DNA if they are not detoxified by phase II enzymes.catechol-O-methyltransferase (COMT)is a critical phase II enzyme, which methylate catechol estrogens. If the methylation reaction is incomplete, these catechol estrogens will be oxidized to semiquinones and quinines, produce reactive oxygen species, which causes DNA damage and tumor initiation (Chakravarti et al., 2001).

COMT gene is presenton chromosome 22q11.2, contains six exons, and expresses at high levels in many tissues including the liver, kidney, breast and endometrium.The COMT Val158Met (rs4680, G--> A) polymorphism has been identified in coding region of protein (Armando et al.,2012), leads to the sustitution of valine (Val) with methionine (Met) at codon 158. This substitution reduced the enzyme activity, individuals with the Met/Met genotype have a 3 to 4 folds lower enzyme activity than those with wild-type Val/Val genotype (Lotta et al., 1995). The frequency of the mutant Met (A) allele vary greatly among the different populations studied, frequency of Met allele is reported as 0.56 in American, 0.5 in European, and 0.27 in Asian populations (Kumar et al.,2017). Since COMT Val158Met polymorphism can reduce the enzymatic activity and may consequently increasethe concentration of circulating carcinogenic catechol estrogens, COMT Va158Met polymorphism is reported a risk factor for prostate cancer initiation which is estrogen dependent. Val158Met has long been the focus of hormone-related cancers such as breast (Lajjin et al., 2013), endometrial (Ashton et al., 2006) and ovary (Delort et al.,2008) cancer etc.

An association between the functional Val158/108Met polymorphism of the COMT gene and prostate cancerhas been investigated in a number of studies,but with contradictory results due to small sample size and different background of included subjects. The aim of the present study was to find out association between COMT Val158Met polymorphism and prostate cancer risk by meta-analysis.

## Method

Meta-analysis was carried out according tometa-analysis of observational studies in epidemiology (MOOSE) guidelines (Stroup et al.,2000).

### Retrieval strategy and selection criteria

Published studies were retrieved through Pubmed, ScienceDirect, Springer Link and Google Scholar databases up to December 31,2019, using followingkey words: ‘Catecho-O-methyltransferase’ or ‘COMT’ or ‘Val158Met’ and ‘Prostate Cancer’ or ‘Cancer. References of retrieved articleswere searched for other eligible articles.

### Inclusion and exclusion criteria

Studies were included if they met the following criteria: (1) investigate the association between COMT Val158Met polymorphism and prostate cancer risk, (2) included studies investigated cases of all types of prostate cancers ie adenoma, squamous and transitionaetc(3)studies with complete information of COMT Val158Met genotype/ allele numbers in prostate cancer cases and controls and (4) sufficient information for calculating the odds ratio (OR) with 95% confidence interval (CI). Major reasons for studies exclusion were as follows: (1) no prostate cancer cases analyzed, (2) the Val158Met polymorphism distribution information missing, and (3) duplicate article.

### Data extraction

The following information was extracted for each eligible study using standard protocols: (i)first author’s family name, (ii)country name, (iii)ethnicity, (iv) year of publication, (v) journal name, (vi) number of cases and controls, (vii)number of genotypes/alleles in cases and controlsand (viii) genotyping method etc. Number of alleles or genotypes in both cases and controls were extracted or calculated from published data to recalculate ORs.

### Statistical Analysis

Pooled ORs with 95 % confidence intervals (CIs) werecalculated using five genetic models: the allele model (Avs. G), the dominant model (AA+AG vs. GG), the homozygote model (AA vs. GG), co-dominant/heterozygote model (AG vs. GG) and the recessive model (AA vs. AG+GG). Heterogeneity was investigated using Q testand quantified by I^2^ statistic (Higgins and Thompson 2002). Both fixed effect and random effect models were used to calculate ORs with their95 % CIs (Mantel and Haenszel, 1959; DerSimonian and Laird 1986), but model adopted on the basis of heterogeneity.in this meta-analysis. The distribution of the genotypes in the control groups were examined for Hardy–Weinberg equilibrium (HWE) using the Chi-square test. Sensitivity analysis wasperformed by removing studies not in HWE from the meta-analysis.Subgroup analysis was conducted byethnicity. Funnel plots, Begg’s and Egger’s testwere used to assess possible publication bias (Begg and Mazumdar,1994; Egger et al.1997). All the P values were two sided, and P value <0.05 was considered statistically significant.All statistical analyses were done using MIX (Bax et al.,2006) and MetaAnalyst (Wallace et al.,2013) program.

## Results

Fifty-three articles have been extracted from the databasesdescribed above. According to inclusion and exclusion criteria,42 articles were excluded, not suitable for inclusion in present meta-analysis(review, comments, meta-analysis, other disease analysed, unrelated to prostate cancer etc.).Selection of study details is shown in Figure 1. Total 10 individual case-control studies were found suitable for inclusion into meta-analysis (Suzuki et al., 2003; Nock et al.,2006; Low et al., 2006; Tanaka et al., 2006; Suzuki et al., 2007; Cussenot et al., 2007; Omrani et al., 2009; Pazarbasi et al., 2013; Brureau et al., 2016; Tang et al., 2017). One author (Brureau et al., 2016) investigated prostate cancer patients from two populations (African and Caucasian), we considered both the samples as separate study. Hence total eleven studies were included in the present meta-analysis.

**Figure 1.**
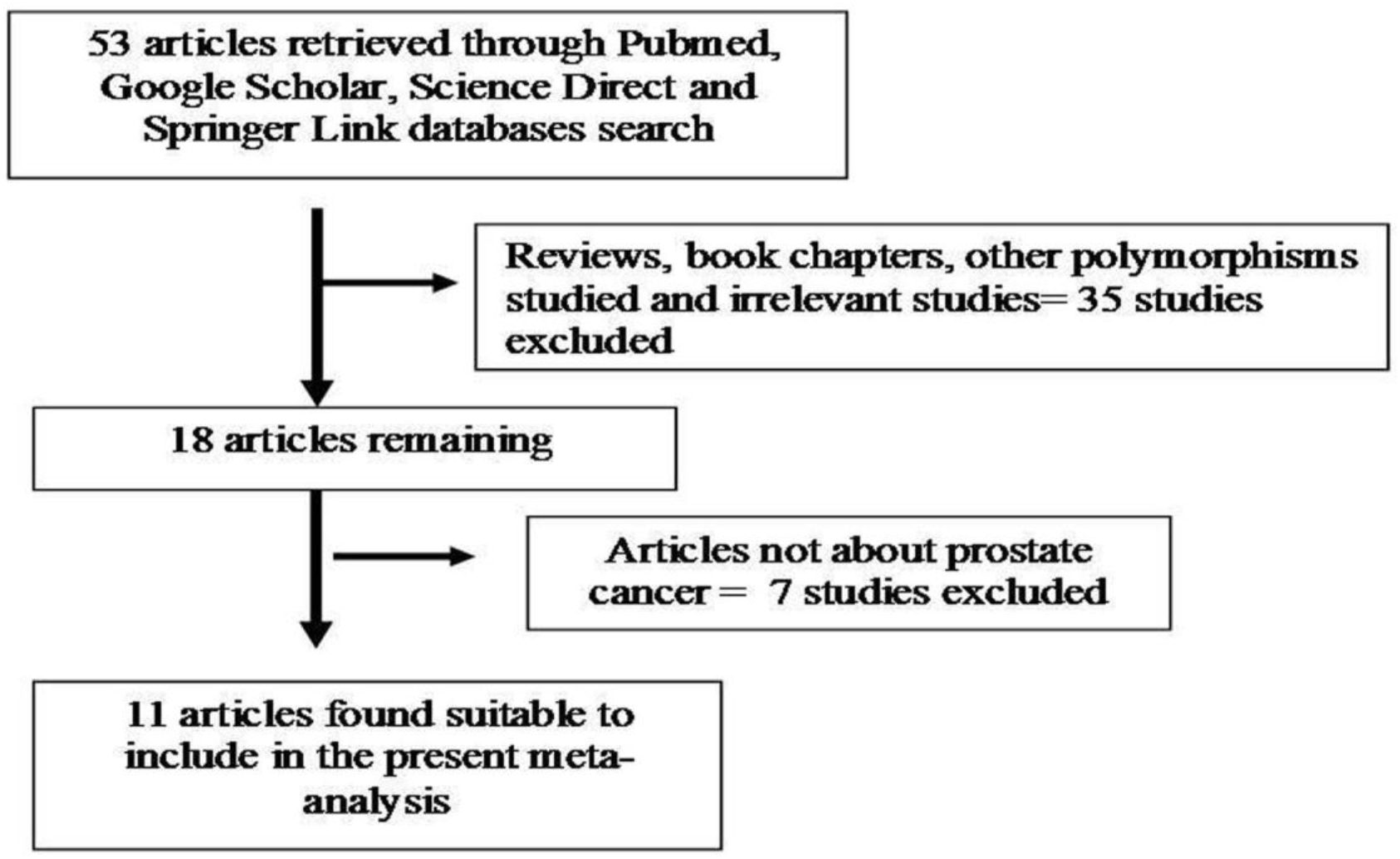
Flow diagram of study search and selection.

In eleven included studies, the smallest case sample size was 34 (Pazarbasi et al., 2013) and largest sample size was 1034 (Cussenot et al., 2007). Total cases and controls were 3,381 and 3,276 respectively. In controls genotypes, percentage of GG, AG and AA were 31.75%, 49.60%, and 18.657%respectively. In total cases, genotype percentage of GG, AG and AA was 34.13%, 49.01% and 16.86% respectively (Table 1). These studies were performed in different countries-France (Cussenot et al., 2007), Iran (Omrani et al., 2009), Japan (Suzuki et al., 2003,2007; Tanaka et al., 2006), Turkey (Pazarbasi et al., 2013), UK (Low et al., 2006), USA (Nock et al.,2006;Brureau et al., 2016; Tang et al., 2017)

**Table 1.**
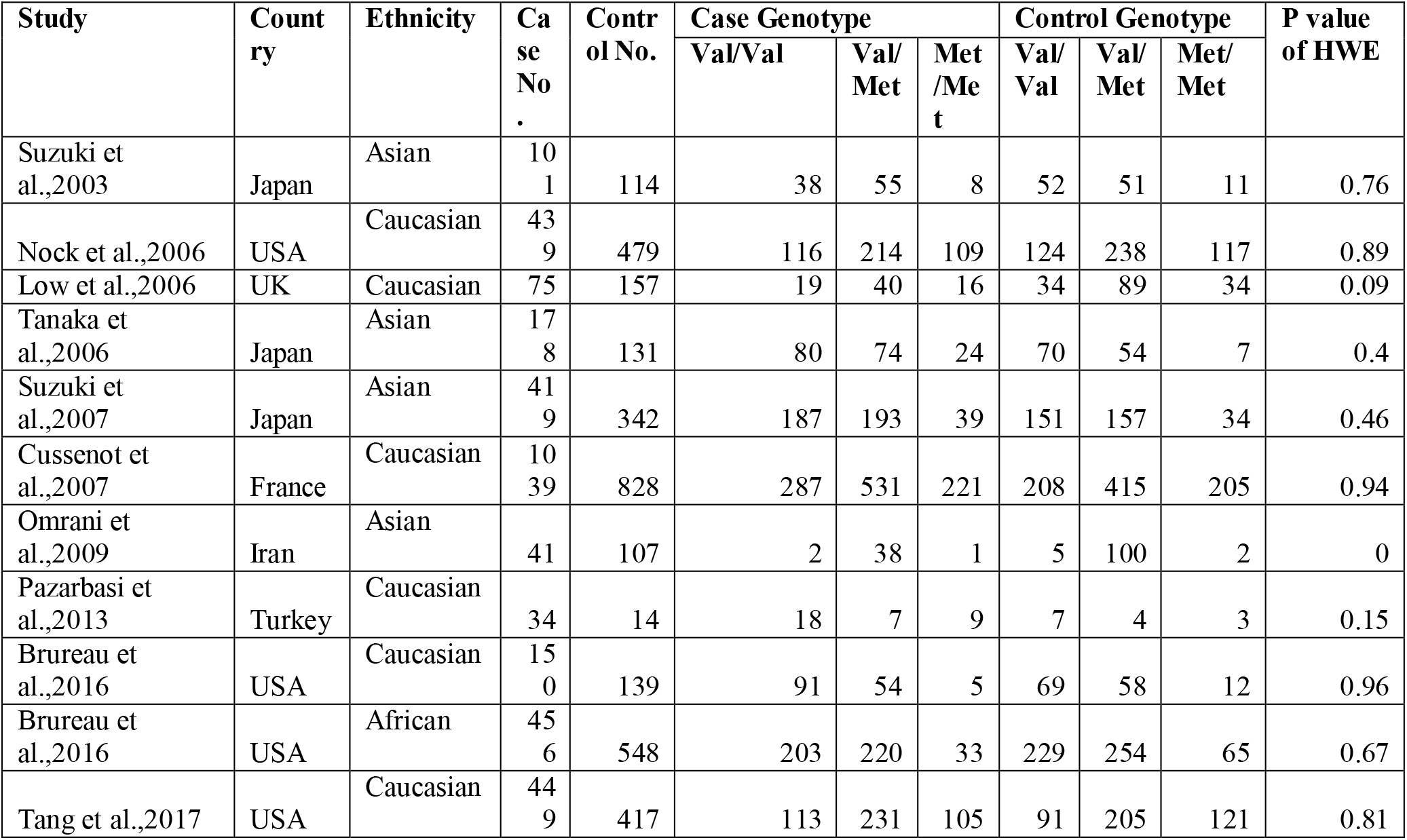
Details of eleven studiesincluded in meta-analysis

### Meta-analysis

Table 2 summarizes the odds ratio with corresponding 95% Confidence intervals for association between COMT Val158Met polymorphism and risk of prostate cancer in allele contrast, dominant, recessive, homozygote and co-dominant models. Meta-analysis with allele contrast did not show significant association between 158Met allele and prostate cancer with both fixed effect (OR_AvsG_= 0.92; 95% CI 0.85-0.98=; p= 0.02) and random effect model (OR_AvsG_=0.93; 95% CI= 0.84-1.02; p=) (Table 1).Co-dominant (OR_AGvs.GG_=0.81; 95% CI=0.85-1.07; p= 0.46) and homozygote (OR_AAvs.GG_= 0.81; 95% CI= 0.70-0.95, p= 0.008) genetic models also did not show any association between COMT Val158Met polymorphism and prostate cancer (Table 2). An increased significant association was found between prostate cancer and dominant model (AA+AG vs. GG) with fixedeffect model (OR_AA+AGvs.GG_ 1.18=; 95% CI= 1.03-1.34; p= 0.01) (Table 2; Figure 2). Association of COMT Val158Met recessivegenotype (AA vs. AG+ GG) was observed significant with prostate cancer using fixed (OR_AAvsAG+GG_= 1.5; 95% CI= 1.31-1.68; p <0.0001) and random (OR_AAvsAG+GG_= 1.54; 95% CI= 1.15-2.07; p = 0.003) effect models (Table 2; Figure 3).

**Table 2.** Summary estimates for the odds ratio (OR) in various allele/genotype contrasts, the significance level (p value) of heterogeneity test (Q test), and the I^2^ metric: overall analysis

| Genetic Models | Fixed effect OR (95% CI), p | Random effect OR (95% CI), p | Heterogeneity p-value (Q test) | I<sup>2</sup> (%) | Publication Bias (p of Egger's test) |
| --- | --- | --- | --- | --- | --- |
| **Total studies** |  |  |  |  |  |
| Allele Contrast (A vs. G) | 0.92(0.85-0.98),0.02 | 0.93(0.84-1.02),0.13 | 0.13 | 32.96 | 0.38 |
| Co-dominant (AG vs. GG) | 0.96(0.85-1.07),0.46 | 0.96(0.85-1.07),0.74 | 0.86 | 0 | 0.98 |
| Homozygote (AA vs. GG) | 0.81(0.70-0.95),0.008 | 0.83(0.66-1.03),0.10 | 0.10 | 36.69 | 0.55 |
| Dominant (AA+AG vs. GG) | 1.18(1.03-1.34),0.01 | 1.17(0.96-1.41),0.11 | 0.12 | 34.65 | 0.59 |
| Recessive (AA vs. GG+AG) | 1.5(1.31-1.68),<0.0001 | 1.54(1.15-2.07),0.003 | <0.0001 | 72.02 | 0.71 |
| **Asian studies** |  |  |  |  |  |
| Allele Contrast (A vs. G) | 1.1(0.93-1.28),0.26 | 1.12(0.91-1.36),0.27 | 0.24 | 28.21 | 0.54 |
| Co-dominant (AG vs. GG) | 1.11(0.88-1.39),0.38 | 1.16(0.87-1.39),0.38 | 0.65 | 0 | 0.61 |
| Homozygote (AA vs. GG) | 1.22(0.82-1.81),0.22 | 1.311(0.70-2.45),0.39 | 0.16 | 41.5 | 0.64 |
| Dominant (AA+AG vs. GG) | 0.86(0.69-1.24),0.42 | 0.81(0.45-1.48),0.50 | 0.16 | 41.82 | 0.68 |
| Recessive ( AA vs. GG+AG) | 2.58(1.79-3.71),<0.0001 | 2.4(1.04-5.52),0.03 | 0.02 | 68.6 | 0.82 |
| **Caucasian studies** |  |  |  |  |  |
| Allele Contrast (A vs. G) | 0.88(0.81-0.96),0.007 | 0.88(0.81-0.96),0.007 | 0.44 | 0 | 0.72 |
| Co-dominant (AG vs. GG) | 0.89(0.77-1.04),0.15 | 0.90(0.77-1.04),0.15 | 0.91 | 0 | 0.12 |
| Homozygote (AA vs. GG) | 0.79(0.66-0.94),0.01 | 0.79(0.66-0.94),0.01 | 0.42 | 0 | 0.65 |
| Dominant (AA+AG vs. GG) | 1.18(1.02-1.37),0.02 | 1.18(1.01-1.37),0.02 | 0.39 | 2.76 | 0.84 |
| Recessive ( AA vs. GG+AG) | 1.44(1.25-1.65),<0.0001 | 1.37(1.01-1.85),0.04 | 0.01 | 66.46 | 0.93 |

**Figure 2.**
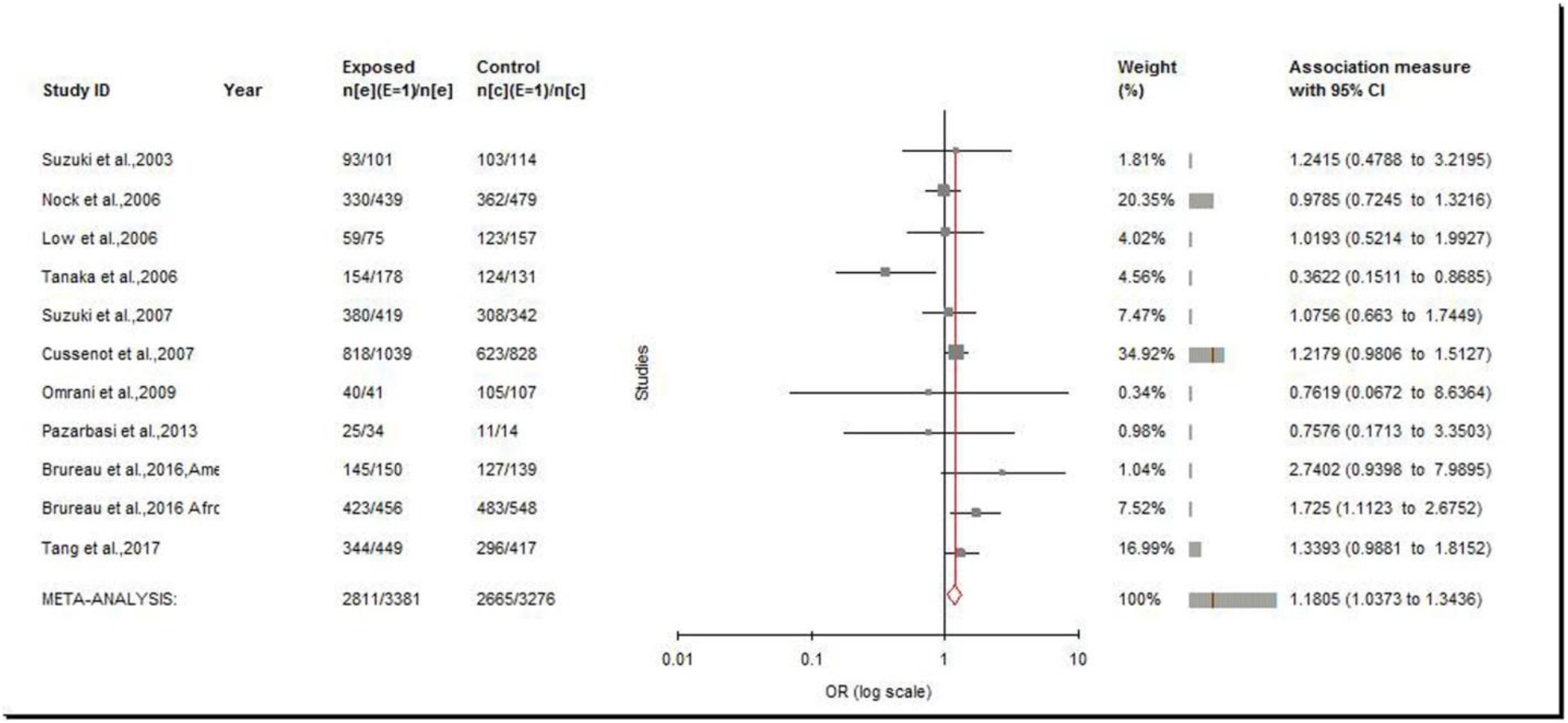
Random effect Forest plot of dominant model (AA +AG vs. GG) of total 11 studies of COMT Val158Met(G472A)polymorphism.

**Figure 3.**
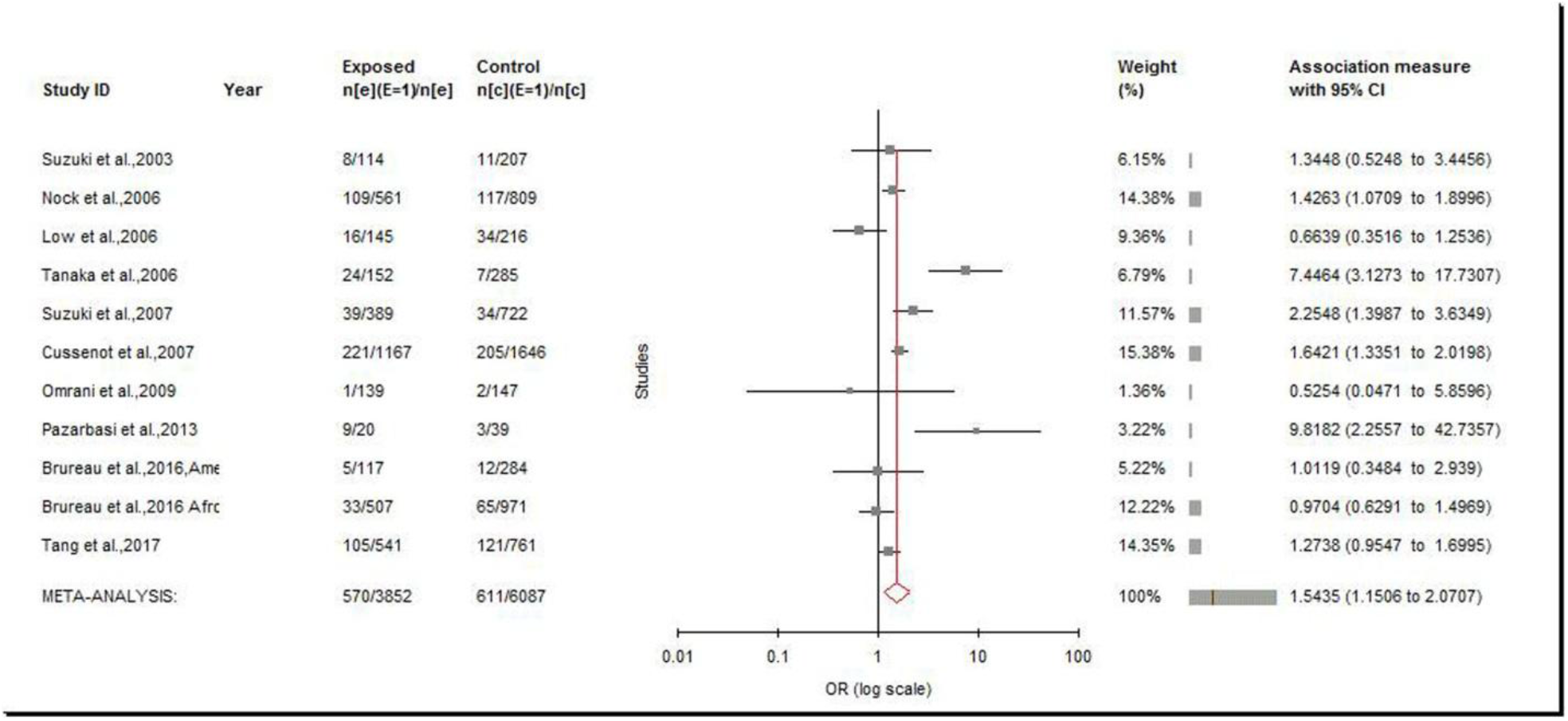
Random effect Forest plot of recessive model (AA vs. AG + GG) of total 11 studies of COMT Val158Met(G472A)polymorphism.

### Heterogeneity and Sensitivity analysis

A true heterogeneity existed between studies for allele contrast (P_heterogeneity_= 0.13, Q= 14.91, I^2^= 32.96%, t^2^= 0.007), homozygote(P_heterogeneity_= 0.10, Q= 15.79, I^2^= 36.69%, t^2^= 0.044), co-dominant (P_heterogeneity_= 0.86, Q= 5.34, I^2^= 0%, t^2^= 0), dominant (P_heterogeneity_= 0.12, Q= 15.30, I^2^= 34.65%, t^2^= 0.03) and recessive (P_heterogeneity_=<0.0001, Q= 35.73, I^2^= 72.02%, t^2^= 0.13) models. Heterogeneity was observed higher only in recessive model.

Sensitivity analysis was performed by eliminating studies with control population deviating from HWE. Control population of one study was not in HWE (Omrani et al.,2009), hence meta-analysis using recessive genetic modelwas performed after eliminating this study,but heterogeneity did not decrease after exclusion of this study.

### Subgroup analysis

Sub-group analysis based on ethnicity was performed. Out of 11included studies, 4 studies were from Asian population, 6studies were from Caucasian populationand 1 study was from African population. In Asian population (number of studies= 4; 739 cases/ 694 controls), allele contrast (OR_AvsG_= 1.1; 95% CI= 0..93-1.28; p= 0.26), co-dominant (OR_AG vsGG_= 1.11; 95% CI= 0.88-1.39; p= 0.38), homozygote (OR_AA vsGG_= 1.22; 95% CI= 0.82-1.81; p= 0.22) and dominant models (OR_AA+AG vs GG_= 0.86 ; 95% CI=0.69-1.24; p= 0.42) did not show any association between COMT Val158Met polymorphism and prostate cancer using both fixed and random effect models (Table 2). Recessive model meta-analysis showed strong statistical association between COMT Val158MEt polymorphism and prostate cancer risk using both fixed effect (OR_AA vs AG+GG_= 2.58; 95% CI= 1.79-3.71; p<0.0001) and random effect(OR_AA vs AG+GG_= 2.4; 95% CI= 1.04-5.52; p= 0.03) models (Table 2; Figure 4).In Caucasian population (number of studies= 6; 2,186 cases/2,034 controls), allele contrast, homozygote and co-dominant meta-analysis did not show association between COMT Val158Met polymorphism and prostate cancer risk (Table 2).Meta-analysis using dominant model showed significant association with both the fixed effect (OR_AA+AGvsGG_= 1.18; 95%CI=1.02-1.37; p= 0.02) and random effect (OR_AA+AG vsGG_= 1.18; 95%CI=1.01-1.37; p= 0.02) models. The recessive model meta-analysis also showed statistically significant association with fixed effect (OR_AA vsAG+GG_= 1.44; 95%CI= 1.25-1.65; p<0.0001) and random effect(OR_AA vs AG+GG_= 1.37; 95% CI= 1.01-1.85; p= 0.04) models(Table 2; Figure 5).

**Figure 4.**
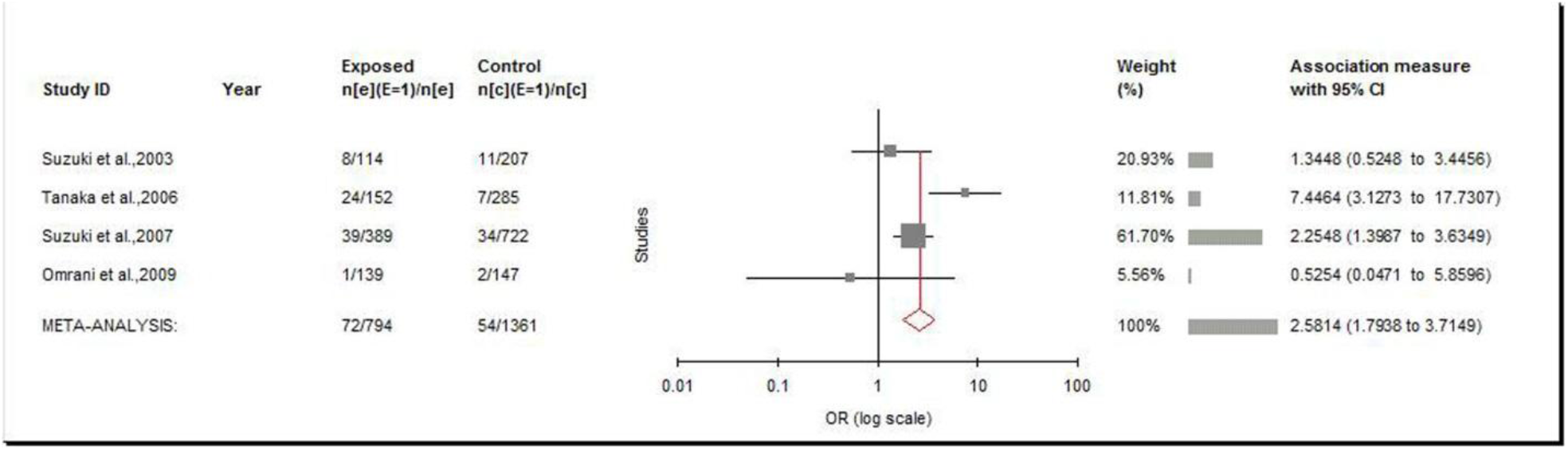
Random effect Forest plot of recessive model (AA vs. AG + GG) of 4 Asian studies ofCOMT Val158Met(G472A)polymorphism.

**Figure 5.**
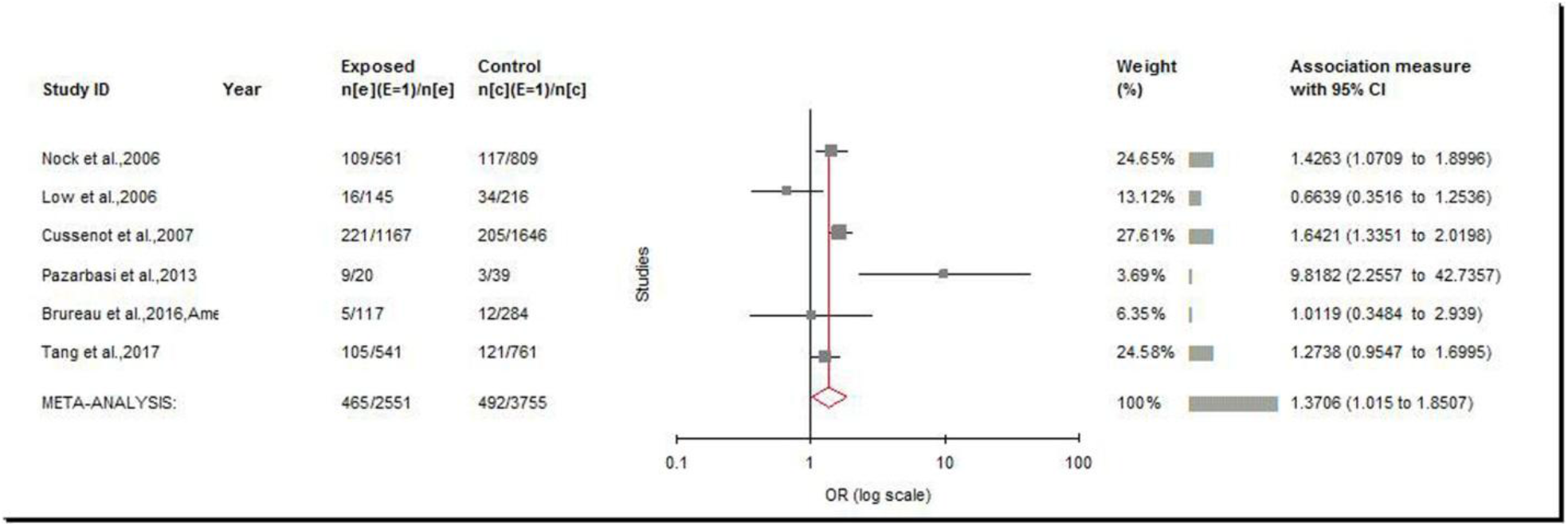
Random effect Forest plot of recessive model (AA vs. AG + GG) of 6 Caucasian studies ofCOMT Val158Met(G472A)polymorphism.

### Publication Bias

Funnel plot observation and p value of Egger’s test showed absence of publication bias in meta-analysis using five genetic models(A vs G, p= 0.38; AA+AG vs GG, p= 0.59; AA vs GG, p= 0.55; AG vs GG, p= 0.98; AA vs. AG+GG, p= 071.) (Table 2; Figures 6,7). Funnel plots of Asian and Caucasian studies also showed absence of publication bias (Table 2)

**Figure 6.**
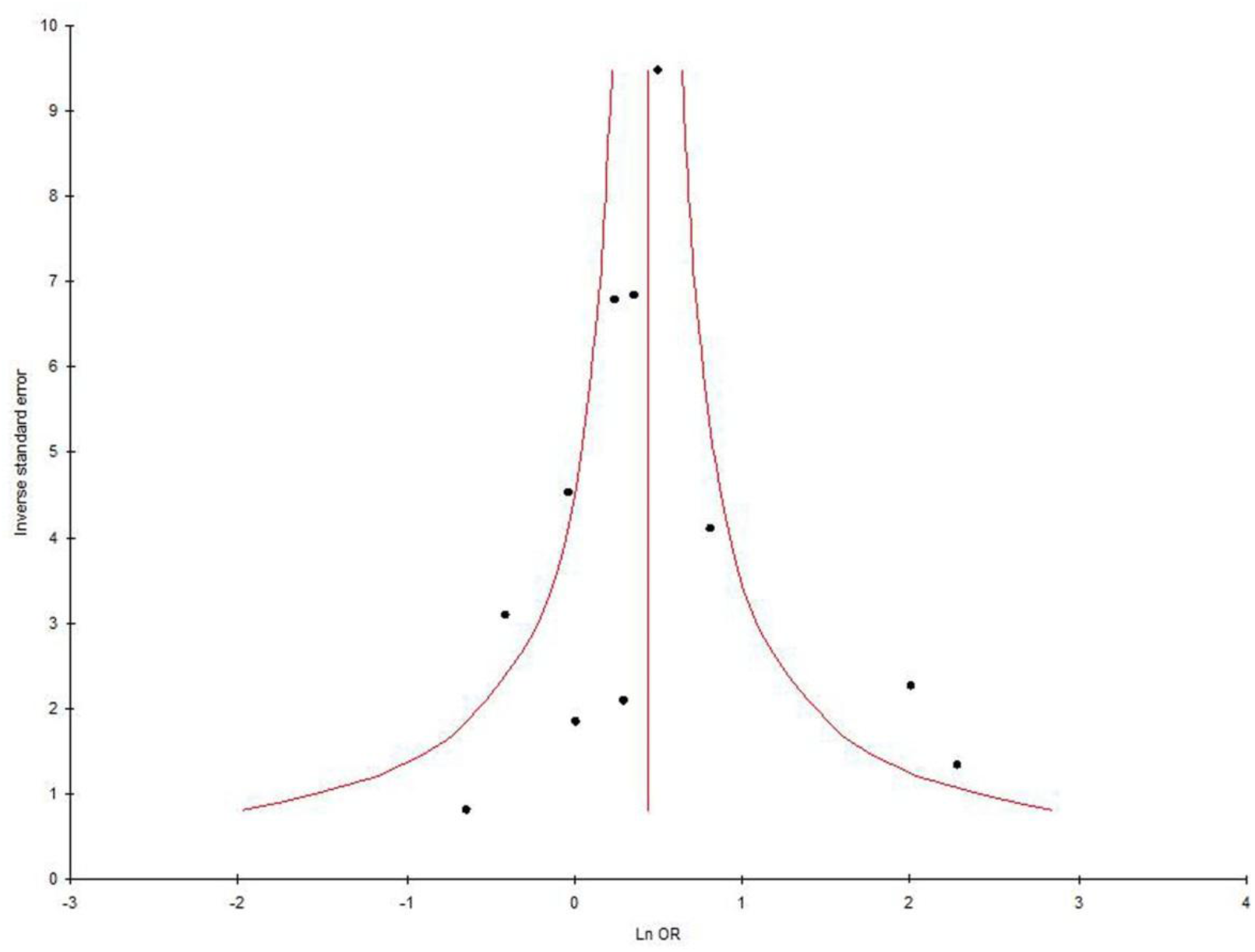
Funnel plot-Precision by log odds ratio for dominant model (AA +AG vs. GG) of total 11 studies of COMT Val158Met(G472A)polymorphism.

**Figure 7.**
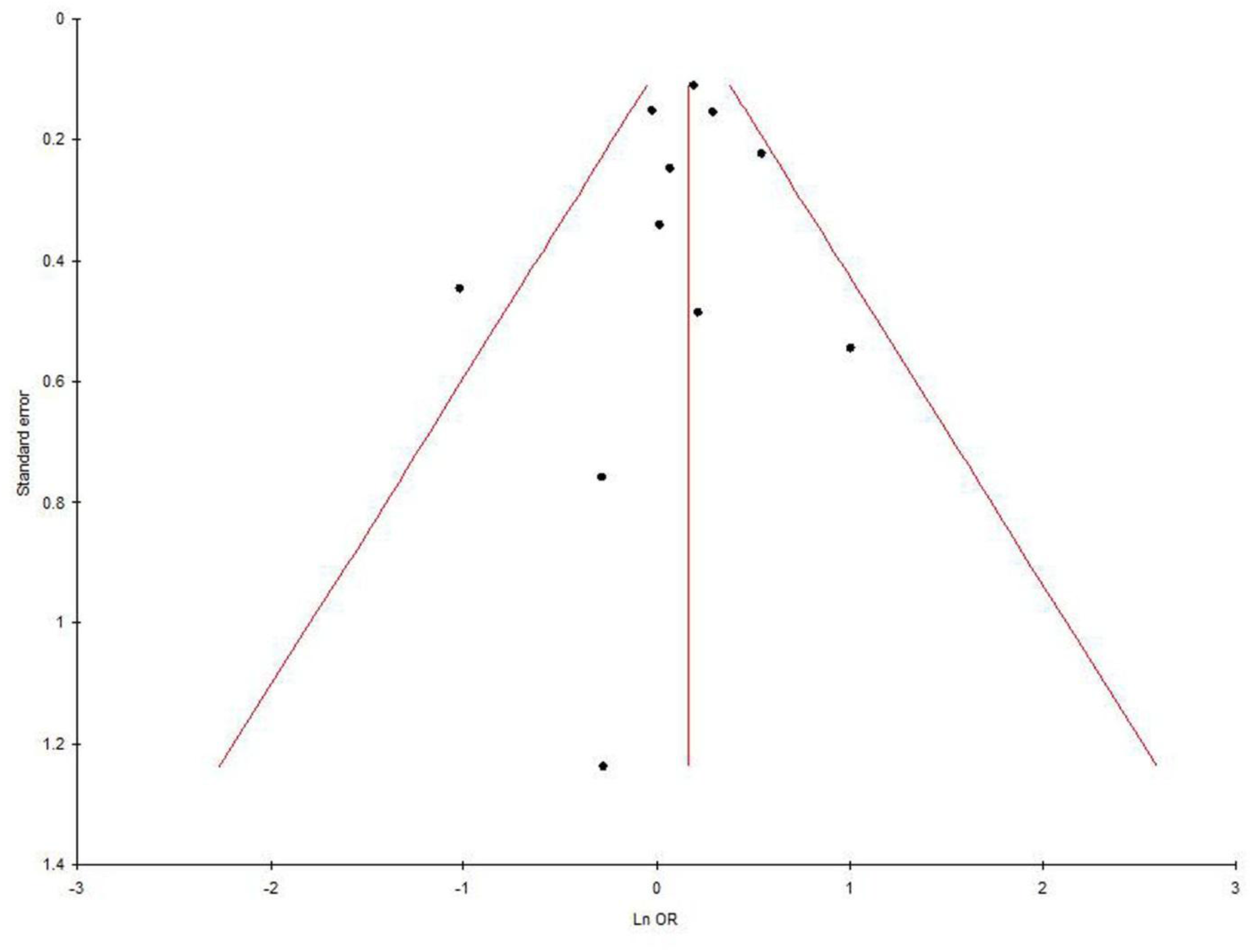
Funnel plot- Standard error by log odds ratio allele contrast model (AA +AG vs. GG) of total 11studies of COMT Val158Met(G472A)polymorphism.

## Discussion

COMT is a phase II enzyme that is involved in the inactivation of catechol estrogens (Axelrod et al., 1958). COMT catalyzes the methylation of catechol estrogens to less polar monomethyl ethers. O-Methylation increases the concentrations of 4-methoxyestradiol (4-MeOE2) and 2-methoxyestradiol (2-MeO-E2) (Dawling et al., 2001). Allelic variation in COMT (Val158Met) is likely related to decrease enzymatic activity and consequently increases the risk of carcinogenesis due to accumulation of estrogen metabolites. Hence, COMT Val158Met polymorphism has been extensively investigated for correlation with different cancer risk like - esophageal cancer (Huang et al.,2011), colorectal cancer (Zhou,2009), hepatocellular carcinoma (Yuan et al.,2008), lung cancer (Zienolddiny et al.,2008), breast cancer (Lajin et al.,2013), ovary cancer (Delort et al.,2008), endometrial cancer (Ashton et al.,2010), testicular germ cell tumor (Ferlin et al., 2010) and bladder cancer (Wolpert et al.,2013) etc.

In presentmeta-analysis, we tried to find out the exact associations between COMT Val158Met polymorphism and prostate cancer susceptibility. Our results indicated that the Val158Met polymorphism is not risk factor for prostate cancer and ORis statistically significant (OR= 1.54; 95% CI= 1.15-2.07; p = 0.003).

Meta-analysis is an acceptable powerful statistical tool, suitable for the dealing with genetic-association data. Mea-analysis overcome deficiencies of small studies/small sample analysis by combining data from several studies and increasing the statistical power (lower type II error rate) (Ioannidis et al., 2003). Several meta-analysis were published which evaluated risk of small effect genes on different disease and disorders-like MTHFR frequency (Yadav et al., 2017), MTRR frequency (Yadav et al., 2019), Down syndrome (Rai,2011; Rai et al., 2017; Rai and Kumar 2018), recurrent pregnancy loss (Rai,2016), cleft lip and palate (Rai,2014,2017), male infertility (Rai and Kumar,2017), obsessive compulsive disorder (Kumar and Rai, 2020), autism (Rai, 2016; Rai and Kumar,2018), epilepsy (Rai and Kumar,2018), schizophrenia (Yadav et al., 2016; Rai et al., 2017), depression (Rai,2014, 2017), Glucose-6-phosphate dehydrogenase deficiency (Kumar et al., 2016), Alzheimers disease (Rai,2016), hyperurecemia (Rai,2016), esophageal cancer (Kumar and Rai, 2018),breast cancer (Rai,2014; Rai et al.,2017), digestive tract cancer (Yadav et al.,2018), colorectal cancer (Rai, 2016), prostate cancer (Yadav et al., 2016), endometrial cancer (Kumar et al., 2020) and ovary cancer (Rai,2016) etc.

Strengths of present meta-analysis include the large sample size and higher statistical power based on large number of cases and controls from differential individual studies. We also performed sensitivity analysis and subgroup analysis.Similar to other meta-analyses, present meta-analysis has also few limitations, which should be acknowledged like-(i) unadjusted OR was used, (ii)control sources were not uniformed, (iii)only four databases were searched for study retrieval, it may be possible that few relevant studies were missed, (iv) single gene polymorphism was considered, (v)other confounding factors such as age, diet, lifestyle, physical activity and environment etc were not considered.

## Conclusions

These results indicate that the COMT Val158Met polymorphism is risk factor for prostate cancer (OR= 1.54; p = 0.003).Subgroup analysis based on ethnicity also confirmed the results that COMT Val158Met polymorphism is risk factor for prostate cancer using recessive genetic model. In future, studies with larger sample sizes from different ethnic population are required to reach a definitive conclusion regarding this association. Also, it is necessary to take into consideration different inheritance patterns and the interaction of the COMT gene with the environment.

## Data Availability

All data are available in the manuscript.

## Abbreviations

COMT: Catechol-O-methyltransferase
Val158Met: Valine158Methionine;
OR: Odd ratio
CI: Confidence interval

## Ethics Approval and consent to participants

In present study, we did not use any human sample, so there is no need of ethics approval and consent of participants.

## Consent for Publication

Yes

## Authors Contributions

VR and PK designed and wrote this manuscript. VR performed the meta-analysis and PK collected data.

